# Comparison of the Mini Parasep SF®, ParaPak SpinCon®, and Paradevice® fecal filtration and concentration devices for microscopic and AI-assisted detection of intestinal parasites

**DOI:** 10.64898/2026.06.02.26354769

**Authors:** Heather Morris, Bobbi S. Pritt

## Abstract

Effective filtration and concentration of stool specimens is an essential pre-analytical step for reducing fecal debris and improving organism recovery using microscopy-based ova and parasite (O&P) examination. This study evaluated three commercially available fecal sedimentation-based filtration/concentration systems – ParaPak SpinCon® (Meridian Bioscience), Mini Parasep SF® (Apacor), and the newly-available Paradevice® (Reingenuity) – for qualitative parasite detection and workflow logistics using conventional and artificial intelligence (AI)-assisted microscopy. Forty clinical stool specimens (20 parasite-positive and 20 parasite-negative) were processed with the 3 devices, and the resultant 120 wet mount and 120 trichrome-stained smear preparations were examined using conventional microscopy. Trichrome-stained slides were also scanned at 40x magnification using a Hamamatsu® NanoZoomerS360® flatbed digital slide scanner and images were analyzed using the Techcyte® Fusion Human Fecal Trichrome AI algorithm. Positive and indeterminate digital findings were confirmed by conventional glass slide microscopy. Slides and digital images were reviewed in a blinded manner. Concordance was assessed among the 360 initial evaluations (microscopy and AI-assisted), and discrepant parasitology results were resolved through re-review and specimen reprocessing as needed. Final qualitative agreement across slide/image evaluations using all three concentration systems was 100%. Minor discrepancies in protozoan and white/red blood cell detection/identification were noted in 5 and 7 cases, respectively, and likely reflected sampling and observer variability. While the three concentration systems produced equivalent qualitative results, the Paradevice® and Mini Parasep SF® offered the most streamlined workflows. These findings support the Paradevice® and Mini Parasep SF® as efficient, analytically equivalent systems that are compatible with traditional and AI-assisted O&P workflows.

**Importance:** Microscopy-based examination of stool specimens remains an important diagnostic tool for intestinal parasitic infections. Filtration and concentration methods improve test performance by increasing organism recovery and reducing obscuring fecal debris prior to evaluation. These methods may also facilitate the preparation of thin monolayer trichrome-stained slides for digital scanning and automated parasite detection. Because many clinical laboratories rely on commercial filtration and concentration systems, supply chain disruptions, product shortages, and manufacturer recalls threaten testing continuity, underscoring the need for validated alternatives. This study demonstrates that multiple commercially available concentration systems provide equivalent parasite detection while supporting both conventional microscopy and emerging digital workflows. These findings provide laboratories with evidence-based alternatives that enhance operational resilience without compromising diagnostic performance.

## Introduction

The stool ova and parasite examination (O&P exam) is an important method for detecting many intestinal parasitic infections (1, 2). The complete O&P exam includes macroscopic inspection of stool for helminths or proglottids and microscopic examination of wet mount and permanently stained preparations for helminth ova, larvae, and protozoa (3) (4). Fecal filtration and concentration are essential preanalytical steps, as they reduce the amount of obscuring fecal debris and increase the likelihood of detecting low-abundance organisms that may be missed in direct smears, respectively (3) (5) (6). The concentrated specimen is used for wet mount preparations and may also be used to prepare permanently stained preparations (7). Concentration is particularly important for AI-assisted detection of protozoan parasites in permanent-stained preparations, as whole-slide imaging and AI-analysis is best performed with thin, debris-reduced stool preparations (8).

In human parasitology, the concentration process usually employs filtration and sedimentation to separate parasites from fecal debris (3). Conventional sedimentation methods, such as the formalin-ethyl acetate procedure, require multiple manual steps including gauze filtration, specimen transfer, and solvent extraction using hazardous chemicals such as ether or ethyl acetate (3). To streamline workflows, increase sensitivity, and reduce exposure to solvents, numerous commercial filtration-based concentration devices have been developed (3). These devices are compatible with commonly used fixatives, including Para-Pak® Ecofix® (Meridian Bioscience, Cincinnati, OH), Apafix®, Alcorfix® (Apacor, Berkshire, UK), TOTAL-FIX® (Medical Chemical Corporation, Torrance, CA) and traditional Zinc-PVA/formalin sets, and eliminate the need for solvent extraction.

Because many clinical laboratories rely on commercial filtration and concentration systems, supply chain disruptions, product shortages, and manufacturer recalls threaten testing continuity, underscoring the need for validated alternatives. This study evaluated three commercially available stool concentration filter devices, including the newly available Paradevice® (Reingenuity, Farmingdale, NY), for qualitative parasite detection using both conventional microscopy and AI-assisted review. Each stool specimen was processed using the three devices, and qualitative detection rates were compared across wet mount, permanent stain, and AI-assisted modalities.

## Materials and Methods

### Clinical Specimens

Forty residual clinical stool specimens previously submitted to the Mayo Clinic Parasitology laboratory for O&P examination were tested, comprising 20 parasite-positive specimens containing a range of commonly encountered protozoa and helminths and 20 parasite-negative specimens, including 5 containing red blood cells (RBCs) and/or white blood cells (WBCs). The specimens were preserved in either 10% formalin/Zinc-PVA collection vial sets or in Ecofix collection vials (see Table 1). Specimens submitted within the past 2 weeks were preferentially selected when available to minimize organism degradation. Each specimen was concurrently processed using all three devices, producing 120 wet preparations and 120 trichrome-stained slides, for a total of 240 slides. This study was approved by the Mayo Clinic institutional review board and biospecimens committee.

**Table 1.**
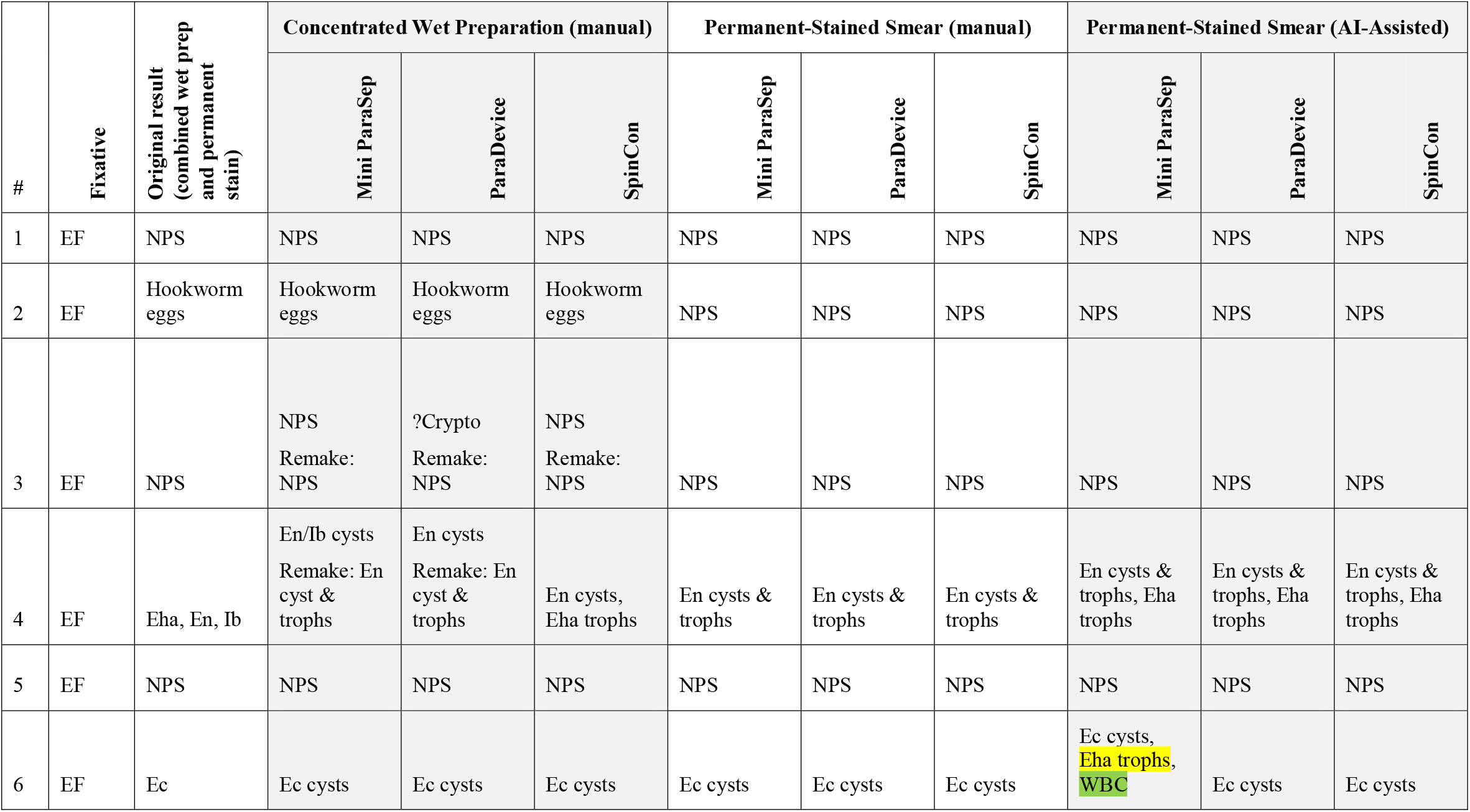
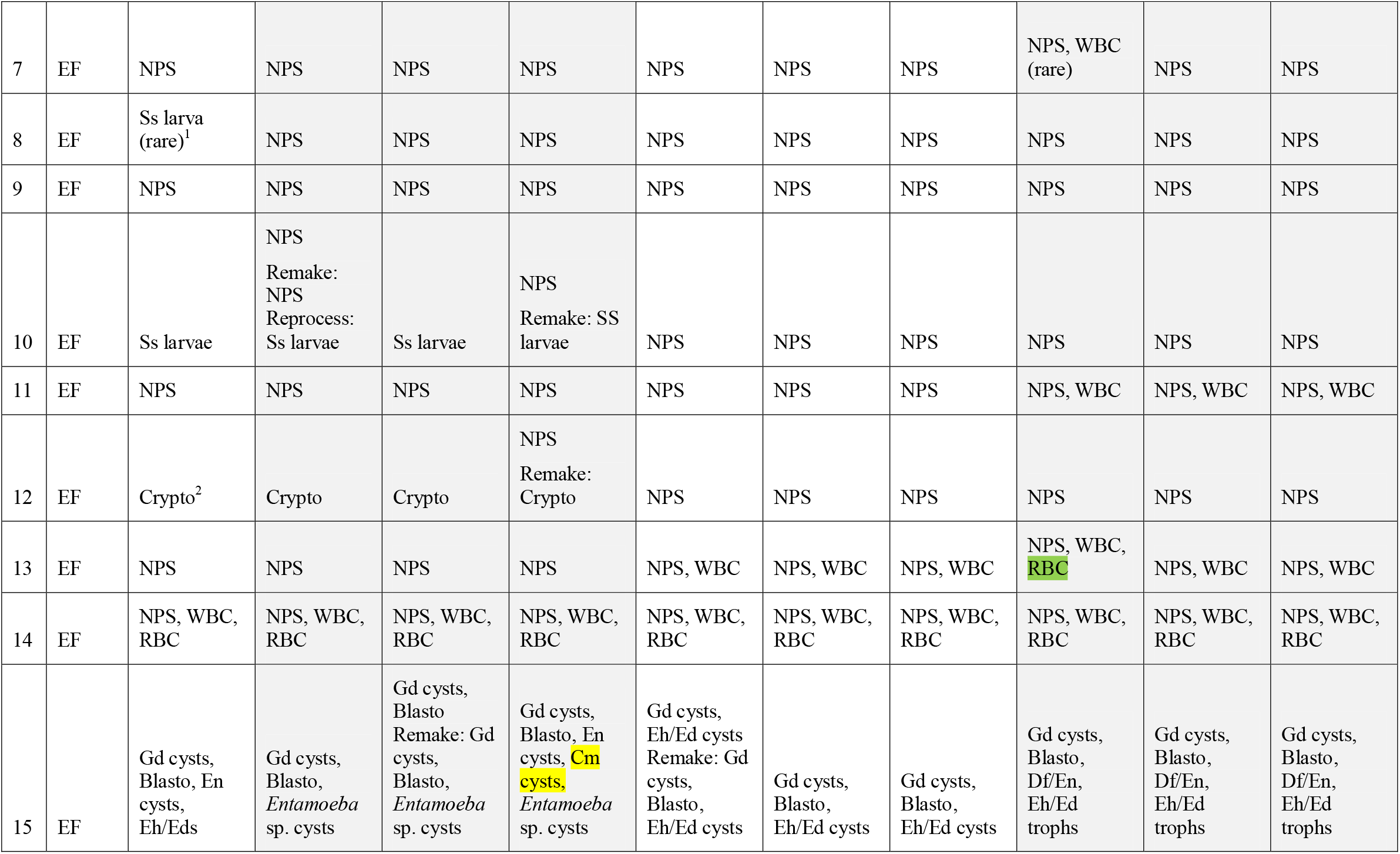
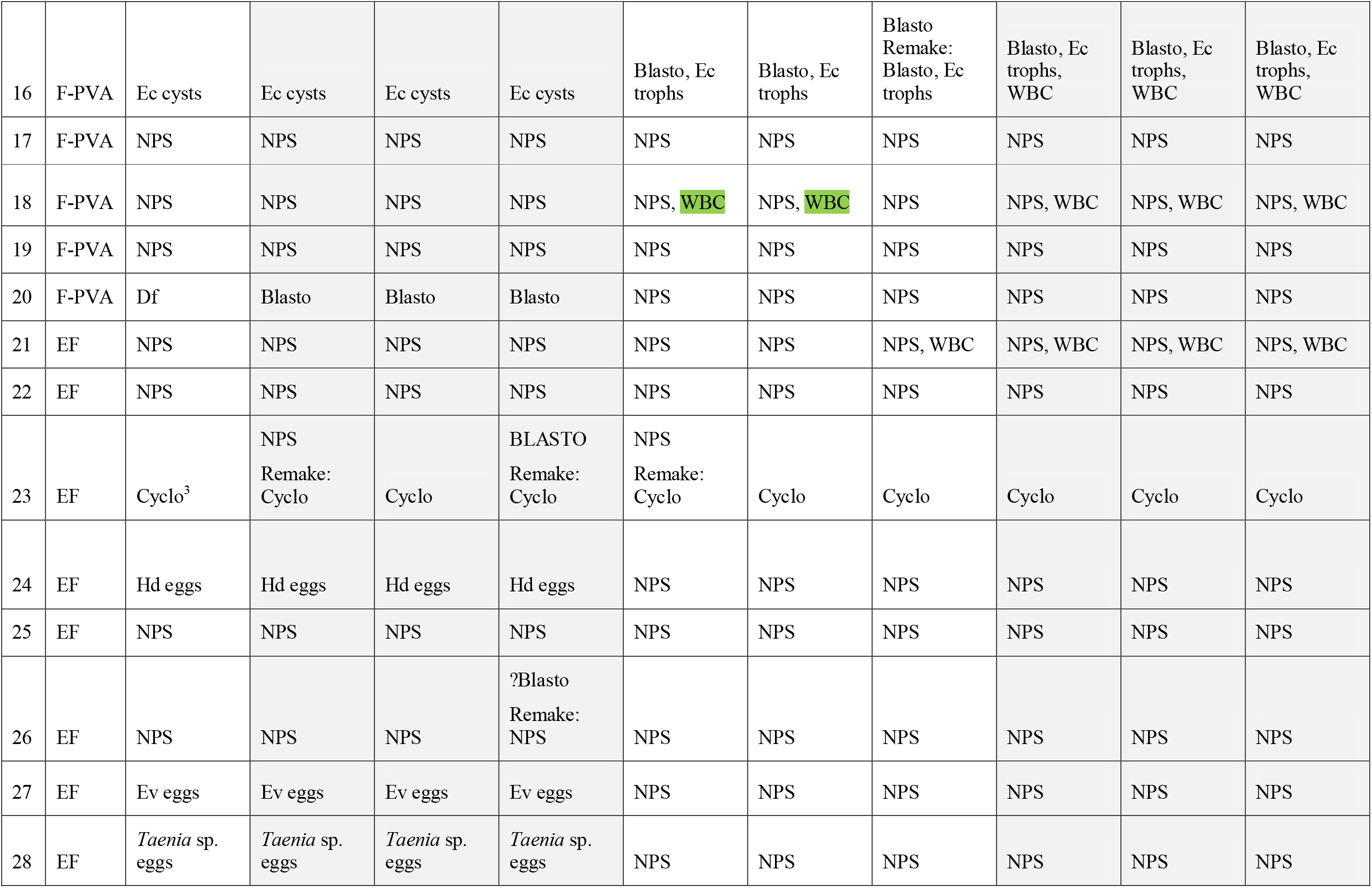
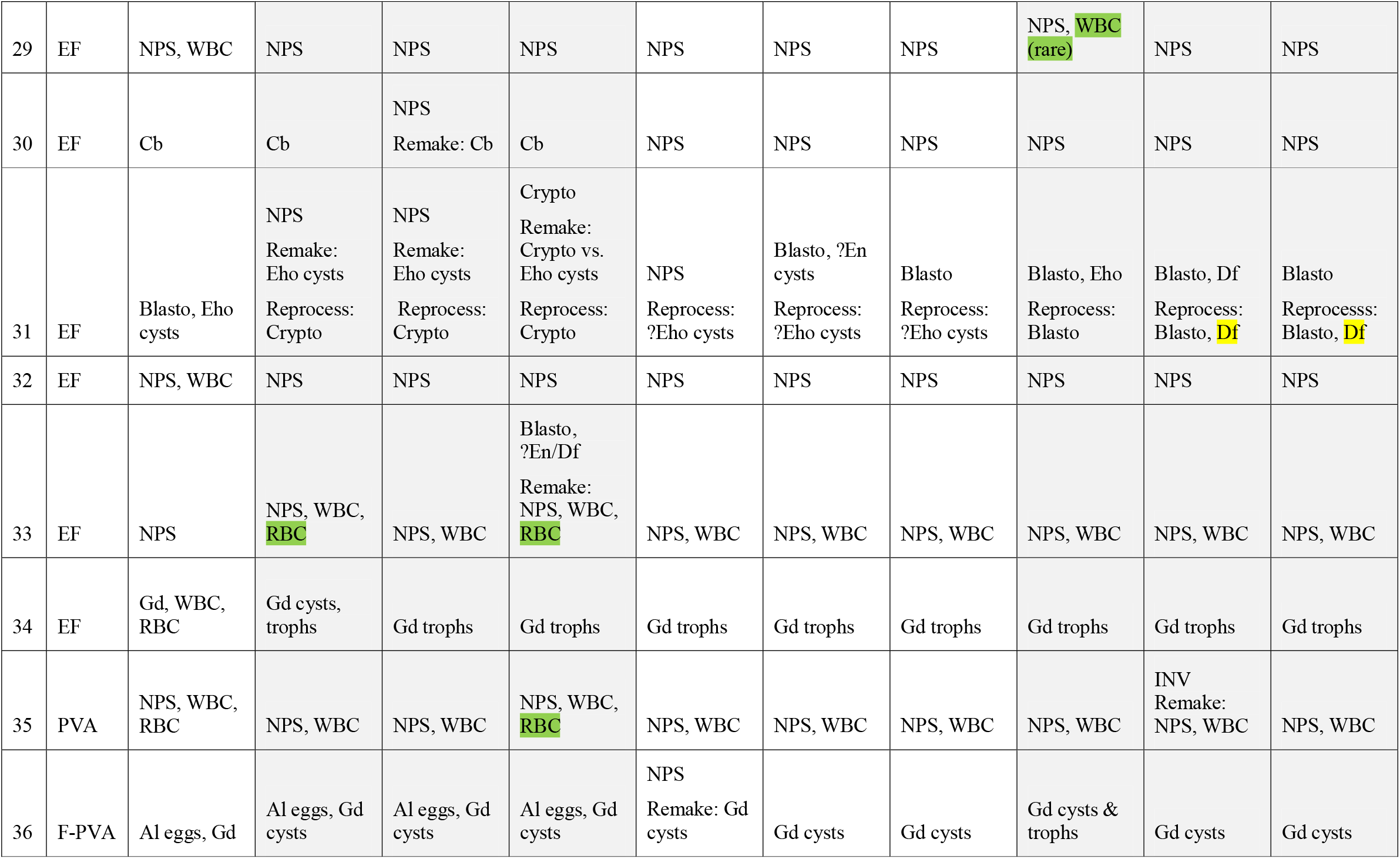
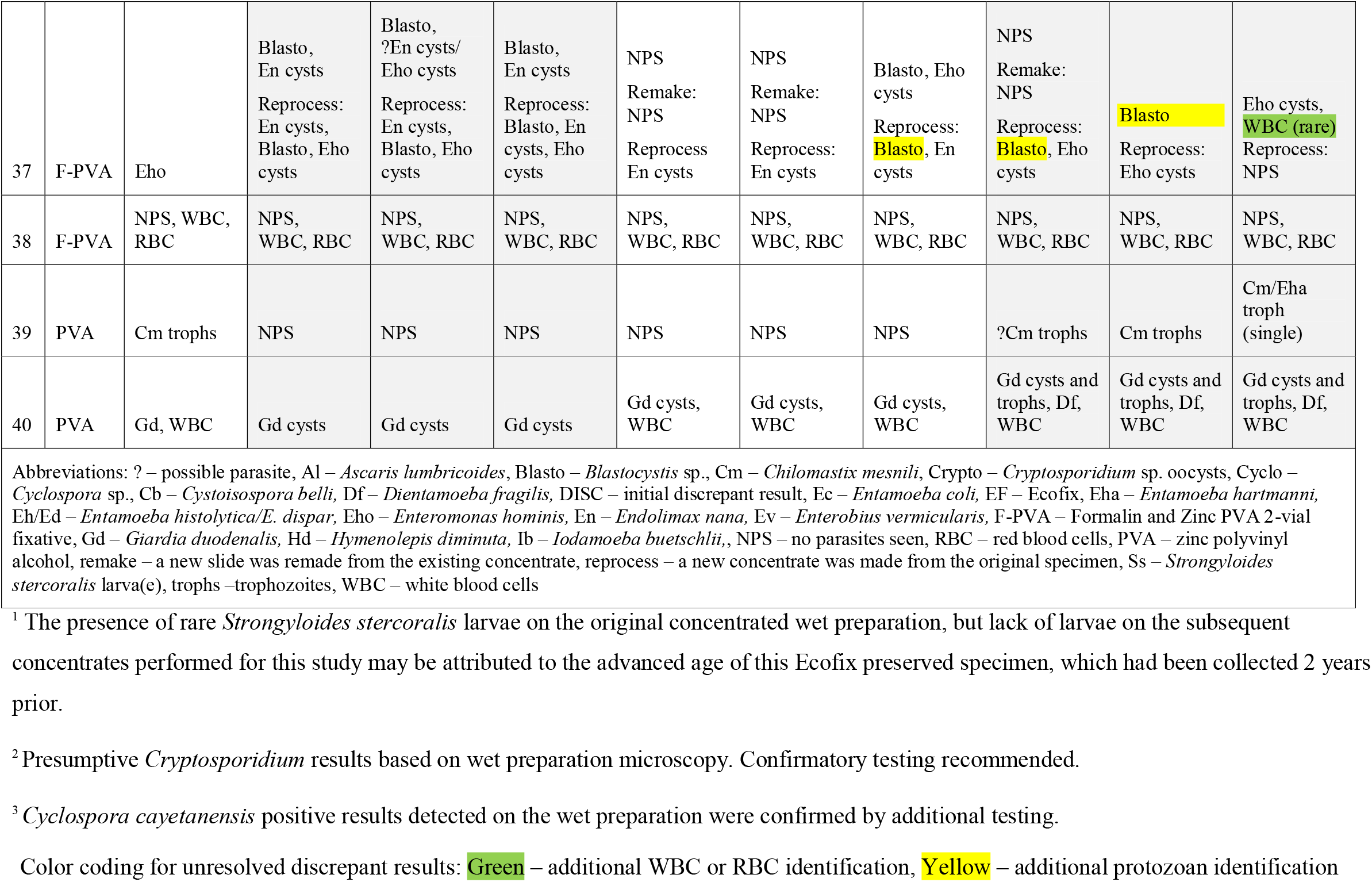
Comparison of results by three concentration systems and type of preparation/examination.

### Concentration methods

Pre-analytic processing steps for each filtration/concentration device are provided in the Appendix. Subsequent processing steps were the same for all 3 concentration devices and are detailed below.

### Examination

Three separate examinations were performed for each concentrate: a wet mount preparation examined by manual microscopic review, a permanent-stained smear examined by manual microscopic review, and the same permanent-stained smear by AI-assisted review after digital imaging. A total of 360 slides/digital images were prepared and reviewed by an experienced technologist in a blinded manner, without knowledge of the original or subsequent results from each concentrate O&P examination. The three preparations were made as follows:

#### Wet mount concentrate preparation

From the reserved fecal pellets set aside after centrifugation during initial specimen processing (see Appendix), one drop of well-mixed stool pellet was placed on a labeled slide using a clean transfer pipette. An additional drop of sterile saline was added to slide if necessary to achieve proper consistency (thin enough to allow microscope light through but still containing as much fecal material as possible). The specimen was covered with a 22 × 22 mm coverslip and examined immediately on a light microscope using 10x and 40x dry objective lenses.

#### Permanent-stained smear preparation with conventional microscopy

One to two drops from the well-mixed fecal pellet were also placed on a separate slide using a clean transfer pipette and spread into a thin, even monolayer with wooden applicator stick on end of a labeled slide. The smear was dried for 30 minutes before staining with Ecostain®, a modified-Trichrome stain protocol (Para-Pak Ecostain®, Meridian Bioscience, Cincinnati, USA) on an automated slide stainer (Leica AutostainerXL, Leica Biosystems, Nussbach, Germany) and permanently mounted with an automated slide coverslipper (Leica Robotic CV5030 Coverslipper, Leica Biosystems, Nussbach, Germany). After drying for 30 minutes, the stained slide was examined on a light microscope equipped using 50x and 100x oil objective lenses.

#### Permanent-stained preparation with AI-assisted review

The same stained slide prepared for manual microscopic exam as described above was used for digital slide imaging and AI-assisted review. Slides were scanned using a Hamamatsu® NanoZoomerS360® flatbed digital slide scanner (Hamamatsu Photonics K.K., Hamamatsu City, Japan) at 40x (dry) magnification with a predetermined scanning profile. Resultant digital slide images were uploaded and analyzed by the Techcyte® Human Fecal Trichrome parasite deep neural network AI algorithm (Techcyte®, Orem, UT). The AI algorithm detects and classifies potential protozoa, white blood cells, and red blood cells based on its locked down algorithm and presents them to the technologist for review and final classification (8). Objects of interest are digitally measured and examined using the same morphologic criteria as with conventional microscopy. All positive or suspected positive slides are confirmed using conventional microscopy.

### Discrepant Analysis

Discrepant results for overall qualitative detection (positive or negative for definitive pathogens) and specific parasite identification (pathogens and non-pathogens) among study specimens were resolved through re-review. If the discrepancy remained, then a new concentrate was made from the original specimen and new slides prepared and examined. For the purposes of this study, definitive pathogens were defined as helminths (eggs, larvae) and the following protozoa: *Giardia duodenalis, Entamoeba histolytica, Cryptosporidium* spp., *Cyclospora* spp., *Cystoisospora belli*. Unless ingested red blood cells were seen, *Entamoeba histolytica* was reported as *Entamoeba histolytica/E. dispar* and was counted as a definitive pathogen despite being unable to make a definitive identification. Given the existing uncertainties of the pathogenic potential of *Dientamoeba fragilis* and *Blastocystis* species, these organisms were not considered definitive pathogens.

## Results

Results of the three-way analysis are shown in Table 1 for all detected parasites, including definitive pathogens and non/uncertain pathogens. Qualitative analysis for definitive pathogens is presented below for each type of preparation.

### Wet Mount Preparation

Initial concordance for pathogenic parasite detection was 87.5% (35/40), with review of wet mounts from from Mini Parasep SF, Paradevice, and SpinCon concentrates missing 2, 1, and 3 definitive pathogens respectively. Following discrepant analysis, concordance reached 100% (40/40) for detection of pathogenic parasites.

### Trichrome Stain with Conventional Microscopy

Initial concordance for pathogenic parasite detection was (38/40), with review of slides from Mini Parasep SF, Paradevice, and SpinCon® concentrates missing 2, 0, and 0 definitive pathogens respectively. Following discrepant analysis, concordance reached 100% (40/40) for detection of pathogenic parasites.

### Trichrome Stain with Digital Imaging and AI-Assisted Examination

Initial concordance for pathogenic parasite detection was 100% (40/40) across examination of slides from from Mini Parasep SF, Paradevice, and SpinCon® concentrates.

### Workflow Observation

The Paradevice® and Mini Parasep SF® devices demonstrated equivalent workflows, characterized by streamlined processing and ease of use. In contrast, the SpinCon system was more labor-intensive, requiring manipulation of multiple components, specimen pre-filtering and rinsing, and additional daily reagent preparation, thereby increasing handling complexity and potential exposure risk.

## Discussion

Effective stool concentration is essential for maximizing parasite recovery and ensuring the most sensitive O&P exam results. This is increasingly important as laboratories adopt digital imaging and AI-assisted diagnostic tools, which require thin, debris-reduced stool slide preparations for optimal outcomes.

In this study, examination of preparations from all three evaluated devices demonstrated equivalent detection of pathogenic parasites. Few definitive parasites were missed on initial examination, and this may have reflected sampling error rather than actual differences in the filtration and concentration efficiency of the three systems. Of note, no definitive pathogens were missed using AI-assisted review of the trichrome stained slides. This is unsurprising given the increased sensitivity of this method (8).

Workflow differences among the three concentration/filtration systems were notable. Paradevice® and Mini Parasep SF® provided the most streamlined, closed-system workflows, reducing hands-on time and minimizing exposure to potentially infectious and hazardous materials. The ParaPak SpinCon® produced acceptable results but required more manual steps and presented more opportunities for accidental spills and exposure.

### Limitations

There are limitations to this study. The comparison was performed in a single laboratory using a relatively small number of parasite-positive and parasite-negative specimens. Examination of additional samples may be informative. Additionally, the examinations were performed by a single, highly experienced technologist with more than 20 years of experience in parasite identification. Lastly, results were limited to a qualitative analysis for definitive parasitic pathogens and did not include semi-quantitative results. Non-pathogens protozoa were not included in the analysis.

## Conclusion

From a diagnostic standpoint, the three filter devices were determined to be comparable in their ability to detect pathogenic parasites and suitable for use in stool O&P testing. Operational differences were noted, with the Paradevice® and Mini Parasep SF® devices offering the most streamlined and efficient processing. Despite workflow distinctions, all three devices provide reliable specimen filtration and concentration for routine laboratory use.

Selection among the evaluated systems may thus be guided by practical considerations such as workflow efficiency, increased staff safety, cost, and supply availability, rather than diagnostic performance. This study provides laboratories with evidence-based alternatives that enhance operational resilience without compromising diagnostic performance.

## Data Availability

All data produced in the present work are contained in the manuscript

## Acknowledgments

Reingenuity® filters were provided at no cost by the manufacturer. The manufacturer had no role in study design, data collection and interpretation, or the decision to submit the work for publication. Dr. Pritt has served as an advisor to GenMark (Roche Diagnostics) and receives royalties from the College of American Pathologists Press.

