## Appendix for "Comparison of the Mini Parasep SF®, ParaPak SpinCon®, and Paradevice® fecal filtration and concentration devices for microscopic and AI-assisted detection of intestinal parasites"

**Appendix: Stool Concentration Filter Device Preparation and Processing Protocols**

Each stool specimen was processed according to the Mayo Clinic Clinical Parasitology Laboratory’s Parasites and Ova in Stool (O&P) standard procedure and the respective manufacturer instructions for each of the three filter devices.  The Mini Parasep SF® (Apacor) and the ParaDevice® (Re Ingenuity) were processed identically due to equivalent design and workflow.  The ParaPak SpinCon® (Meridian) was processed according to an alternate validated protocol and manufacturer instructions, which included two additional steps: 1) use of a separate funnel‑shaped filter component and 2) a saline/surfactant wash to facilitate specimen passage into the centrifugation tube.

Mini Parasep SF® Fecal Parasite Concentrator device:

1. Approximately 2-3 mL of stool specimen was aliquoted into the tube portion of the concentration device with a clean plastic transfer pipette. The concentration device was assembled and sealed by screwing the conical-ended filter unit into the tube portion of the device. The device was inverted, placing the conical end down for centrifugation.
2. The device was then centrifuged at 2000 RPM for 10 minutes, and the supernatant was discarded, reserving the fecal sediment pellet.
3. 0.9% normal saline was added to the fecal pellet in an amount approximately 2 times the volume of pellet and mixed to resuspend the fecal pellet with a clean plastic transfer pipette.
4. If the specimen consisted of a 2-vial set of formalin-preserved stool and Zn-PVA-preserved stool, then 2 concentration devices were prepared: the formalin-preserved fecal pellet to be used for the wet mount portion of the O&P examination and the Zn-PVA preserved fecal pellet to be used to prepare the permanent-stained smear.
5. Subsequent processing steps were the same for all 3 concentration devices and can be found below.

Paradevice® Intestinal Parasite Concentrator device:

Note: The pre-analytic processing steps for the Paradevice® Concentrator are identical to those for the Mini Parasep SF® as described above.

1. Approximately 2-3 mL of stool specimen was aliquoted into the tube portion of the concentration device with a clean plastic transfer pipette. The concentration device was assembled and sealed by screwing the conical-ended filter unit into the tube portion of the device. The device was inverted, placing the conical end down for centrifugation.
2. The device was then centrifuged at 2000 RPM for 10 minutes, and the supernatant was discarded, reserving the fecal sediment pellet.
3. 0.9% normal saline was added to the fecal pellet in an amount approximately 2 times the volume of pellet and mixed to resuspend the fecal pellet with a clean plastic transfer pipette.
4. If the specimen consisted of a 2-vial set of formalin-preserved stool and Zn-PVA-preserved stool, then 2 concentration devices were prepared: the formalin-preserved fecal pellet to be used for the wet mount portion of the O&P examination and the Zn-PVA preserved fecal pellet to be used to prepare the permanent-stained smear
5. Subsequent processing steps were the same for all 3 concentration devices and can be found below.

ParaPak SpinCon® Stool Concentration System Filter Device:

Note: Pre-analytic processing of the ParaPak SpinCon® device included some significant differences between the Mini Parasep SF® concentration filter device and the Paradevice® Intestinal Parasite Concentrator, including 2 additional steps for the SpinCon® concentrator: 1.) the addition of a separate funnel-shaped filter used on the top of the centrifugation tube and 2.) a saline/surfactant solution wash to aid the stool specimen’s passage through this funnel-shaped filter into the centrifugation tube portion of the concentration device below.

1. Approximately 2-3 mL of stool specimen was aliquoted onto filtration device funnel placed atop conical-bottomed filter tube.
2. Approximately 3 mL of previously prepared SpinCon® saline/surfactant solution was pipetted onto filtration device funnel directly on top of stool specimen in order to wash stool through funnel filter.  Once liquid specimen had passed through and mostly solid parts of stool specimen were left on funnel filter surface, the funnel portion of the device was discarded.
3. The conical-ended tube part of the device was capped, shaken vigorously, and inverted, placing the conical end down for centrifugation.
4. The device was then centrifuged at 2000 RPM for 10 minutes, and the supernatant was discarded, reserving the fecal sediment pellet.
5. 0.9% normal saline was added to the fecal pellet in an amount approximately 2 times the volume of pellet and mixed to resuspend the fecal pellet with a clean plastic transfer pipette.
6. If the specimen consisted of a 2-vial set of formalin-preserved stool and Zn-PVA stool, then 2 concentration devices were prepared: the formalin-preserved fecal pellet to be used for the wet mount portion of the O&P examination and the Zn-PVA preserved fecal pellet to be used to prepare the permanent-stained smear.
